# CluSA: Clustering-based Spatial Analysis framework through Graph Neural Network for Chronic Kidney Disease Prediction using Histopathology Images

**DOI:** 10.1101/2022.12.13.22283319

**Authors:** Joonsang Lee, Elisa Warner, Salma Shaikhouni, Markus Bitzer, Matthias Kretzler, Debbie Gipson, Subramaniam Pennathur, Keith Bellovich, Zeenat Bhat, Crystal Gadegbeku, Susan Massengill, Kalyani Perumal, Jharna Saha, Yingbao Yang, Jinghui Luo, Xin Zhang, Laura Mariani, Jeffrey B. Hodgin, Arvind Rao

**Author notes:** Correspondence should be addressed to J.L., J.B.H., A.R.

## Abstract

Machine learning applied to digital pathology has been increasingly used to assess kidney function and diagnose the underlying cause of chronic kidney disease (CKD). We developed a novel computational framework, Clustering-based Spatial Analysis (CluSA), that leverages unsupervised learning to learn spatial relationships between local visual patterns in kidney tissue. This framework minimizes the need for time-consuming and impractical expert annotations. To incorporate spatial information over the clustered image patterns on the biopsy sample, we spatially encoded clustered patterns with colors and performed spatial analysis through graph neural network. A random forest classifier with various groups of features were used to predict CKD. For predicting high/low eGFR at the biopsy, we achieved a sensitivity of 0.97, specificity of 0.90, and accuracy of 0.95. AUC was 0.96. For predicting eGFR changes in one-year, we achieved a sensitivity of 0.83, specificity of 0.85, and accuracy of 0.84. AUC was 0.85. This study presents the first spatial analysis based on unsupervised machine learning algorithms. Without expert annotation, CluSA framework can not only accurately classify and predict the degree of kidney function at the biopsy and in one year, but also identify novel predictors of kidney function and renal prognosis.

## INTRODUCTION

### Background and Significance

Chronic Kidney Disease (CKD) involves a gradual loss of kidney function and is not easily detected in the early stages until the condition is advanced. According to the Centers for Disease Control and Prevention, more than 37 million people (15% of US adults) are estimated to have Chronic Kidney Disease (CKD) and as many as 9 in 10 adults with CKD do not know they have CKD.^1^ Diabetes, high blood pressure, heart disease, and a family history of kidney failure are the most common causes of kidney disease. Currently, CKD, which causes more deaths than breast cancer or prostate cancer, is the 9^th^ leading cause of death in the U.S.^1^

As the degree of kidney dysfunction is associated with increased mortality and risk of heart disease,^2, 3^ an early, accurate diagnosis is crucial to slow the progression to kidney failure.^4^ Current typical measures of kidney function and risk of progression such as creatinine level in the blood and protein in the urine^5, 6^ have several limitations and are not accurate at higher levels of kidney function.^7^ Although kidney biopsy samples may provide further prognostic information, e.g., degree of glomerular sclerosis and interstitial fibrosis,^8^ they are often visually estimated, and interpretation may vary among pathologists. Computer-aided algorithms may provide a more objective kidney assessment and help to overcome substantial inter-observer variability.

Several deep learning and machine learning approaches to histopathological image analysis have become increasingly common with the growing availability of whole-slide digital scanners.^9^ Coudray *et al*. used convolution neural networks (CNN) on whole-slide images (WSI) to classify them into lung adenocarcinoma (LUAD), lung squamous cell carcinoma (LUSC) or normal lung tissue.^10^ Also, CNN have been applied to WSI to classify sclerosed and non-sclerosed glomeruli.^11, 12^ Kolachalama *et al*. demonstrated that CNN deep learning models can outperform the pathologist-estimated fibrosis score across the classification tasks and can be applied to routine renal biopsy images.^13^

To date, most machine learning and deep learning algorithms applied to histopathology images have been based on supervised (training) approaches. However, supervised algorithms require the use of a large amount of labeled training data, which is a time-consuming, often impractical, and expensive task. To overcome this problem, several studies have proposed methods such as weakly-supervised learning and multiple instance learning (MIL) which gave relatively high performance.^14-18^ However, these are still supervised methods that required patient-level labels.

Spatial analysis of tissue microenvironment and cellular organization has become increasingly popular with multiplexed staining and advanced visualization techniques.^19^ Investigating the spatial context in digital histopathology images is key to understanding the microenvironment heterogeneity with clinical implications^20^. In recent years, graph neural networks (GNNs)^21^ have demonstrated ground-breaking performances in various deep learning applications such as graph convolutional network (GCN),^22^ deep graph convolutional network (DGCNN),^23^ and graph attention network (GAT).^24^

The primary objective of this study was to propose the novel computational framework that does not require expert’s annotation and investigated the spatial context of histopathology images through CluSA with GNN. We hypothesize that adding spatial neighborhood information, which is an important characteristic feature of all forms of CKD, to the clustering analysis can help to improve the predictive model performance. The overall computational pipeline of this study is summarized in Figure 1.

**Figure 1.**
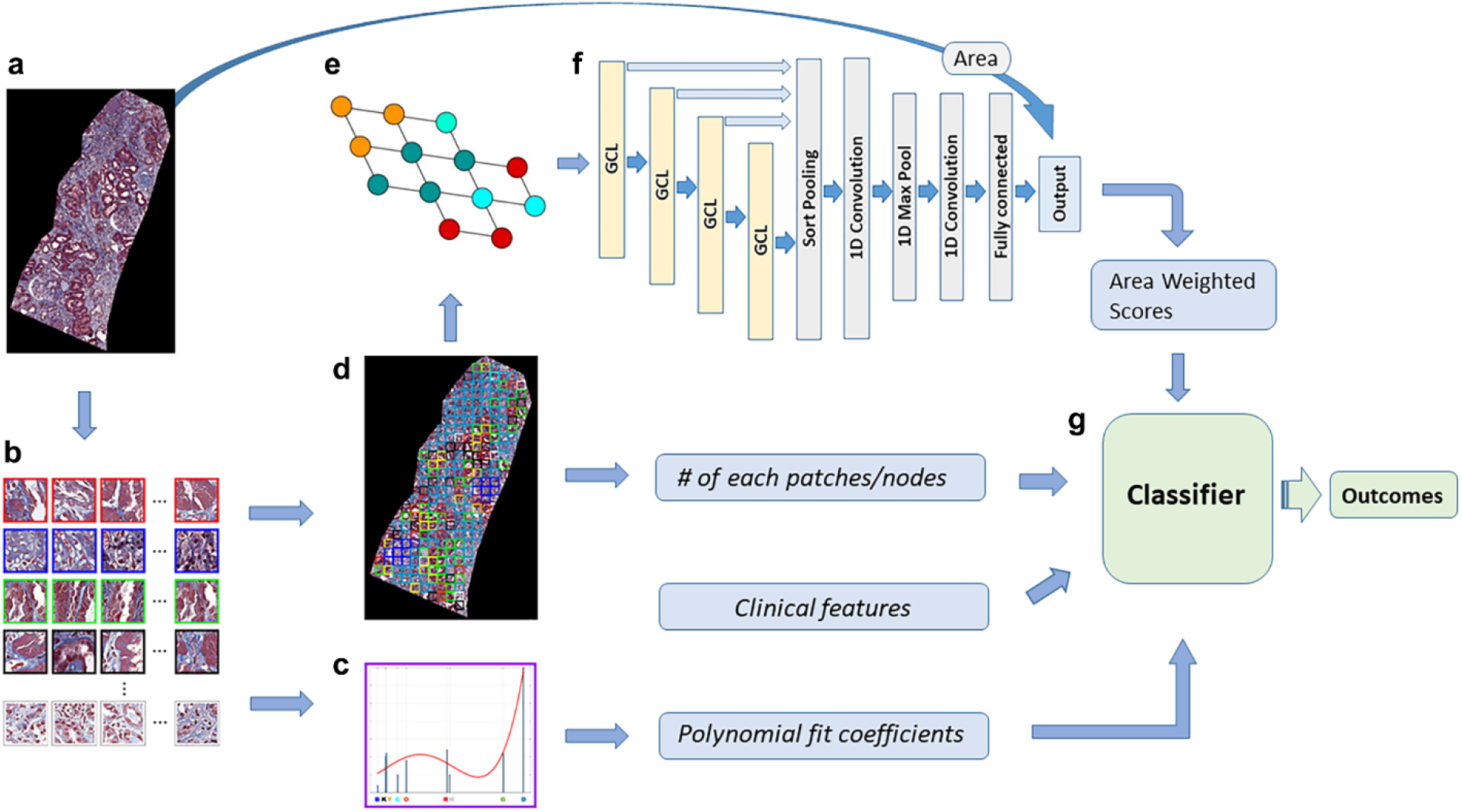
Overall workflow. (a) biopsy core image; (b) clustered patches obtained using a k-means clustering algorithm; (c) histogram of clusters; (d) color-coded map consisting of clustered patches; (e) graph representation converted from the color-coded map; and (f) DGCNN model that outputs core-level prediction. The normalized area weighted score (aw-score) was computed with the area of each core biopsy sample for the patient-level prediction. (g) Random forest machine learning classifier was used with various groups of features, such as aw-scores, the number of clustered patches or nodes, clinical features, and polynomial fit coefficient features.

## MATERIALS AND METHODS

In this study, we proposed a novel computational ensemble machine learning framework, clustering-based spatial analysis (CluSA), that utilizes both unsupervised and supervised machine learning methods to predict patient outcomes as well as to identify important patterns or features associated with the level of kidney function and risk of progression. The unsupervised machine learning using a clustering method learns from an unlabeled dataset and automatically finds structures or patterns in the data by extracting useful features^25^ and the supervised machine learning using graph analysis learns spatial information through neighbor relationships between adjacent structures or patterns in the data.

### Data collection

The dataset consisted of human subjects enrolled in the C-PROBE cohort, a multicenter cohort of patients with CKD established under the auspices of the George O’Brien Kidney Center at the University of Michigan (https://kidneycenter.med.umich.edu/clinical-phenotyping-resource-biobank-core). The C-PROBE aimed at collecting high-quality data and biosamples for translational studies approved by the Institutional Review Boards of the University of Michigan Medical School (IRBMED) with approval number HUM00020938. C-PROBE enrolls patients at the time of clinically indicated biopsy and follows them with phenotypic data prospectively. The written informed consent was obtained from all subjects and/or their legal guardians.

A total of 107,471 histopathology images (256 × 256 pixels) were used in the clustering and in the deep learning model. All images were obtained from 172 biopsy cores from 57 cases in the form of trichrome-stained slides. This study was conducted and carried out in accordance with relevant guidelines and regulations. The Chronic Kidney Disease Epidemiology Collaboration (CKD-EPI) formula was used to calculate the estimated glomerular filtration rate (eGFR).^7, 26^

### Preprocessing

In modern digital pathology, stain normalization is an important processing task for the computer-aided diagnosis (CAD) systems. In this study, we used Reinhard color normalization on all whole-slide-imaging (WSI) data as a preprocessing step to avoid confounding due to variations in color and intensity of the images and increase the computational efficiency and performance.^27^ The Reinhard normalization algorithm maps the color distribution of the stained image to that of a reference image by using a linear transform in a perceptual color space. We computed the global mean and standard deviation of each channel in Lab color space for all data and used them as reference values to normalize our data. Figure S1 in the Supplementary materials provides an example of data with stain normalized images.

### Feature extraction for the clustering using transfer learning method

Feature extraction is an important step in clustering, and it aims at extracting relevant information which characterizes each image pattern. In this process, relevant features are extracted from images to form feature vectors that are used to cluster image patterns based on similarity measures. There are several feature extraction methods such as Histogram of Oriented Gradients (HOG), Scale Invariant Feature Transformation (SIFT), Speeded-Up Robust Features (SURF), and Features from Accelerated Segment Test (FAST).^28, 29^ In this study, for feature extraction, we used one of the most popular machine learning methods called transfer learning.^30-32^ Transfer learning is especially popular in medical image analysis for deep learning where the data are not sufficient for training.^33-35^ Transfer learning uses a pre-trained deep learning model where a model was developed for a task and reused as the starting point for a model on another related task.^36^

We used DeepLab V3+ with ResNet-18 architecture^37, 38^ pre-trained on ImageNet.^39^ We performed feature extraction on this pre-trained deep learning model, allowing the input image to propagate forward and take the outputs of the specific layer as our features. We extracted features from a layer (res5b) of ResNet-18,^40^ a decoder structure, which is a part of deep neural networks for semantic segmentation, DeepLab V3+.

### Clustering and histogram features

We used one of the most popular and simplest unsupervised machine learning algorithms called K-means clustering, which forms groupings using a similarity or distance measures. First, the optimal number of data cluster K was estimated from the Silhouette algorithm in MATLAB (Supplementary Figure S2). Then, the K-means clustering algorithm was applied on the feature vectors for each image tile, obtained from all the images across the patients, to obtain cluster indices, centroid locations, and distances from each point to every centroid. Then, we constructed the histogram representation for each case based on nearest-centroid group labels assigned to each point, resulting in 9 visual pattern group bins for the histogram. The frequency on the cluster histogram represents how often each clustered image pattern is encountered for each case. This occurrence or frequency for each visual pattern was used as a feature for predicting the patient’s eGFR. The Chronic Kidney Disease Epidemiology Collaboration (CKD-EPI) formula was used to calculate eGFR.^7, 26^ The order of clusters on the x-axis of the cluster histogram was obtained using multidimensional scaling (MDS). MDS allows us to visualize how near groups are to each other in histogram plots by computing the similarity between clustered visual pattern groups with the Euclidean distances.

In this study, we used the 4^th^ polynomial fit coefficients as an additional feature for predicting patients’ outcomes. We applied the 4^th^ polynomial fitting on the frequency histogram and obtained five coefficients for each case.

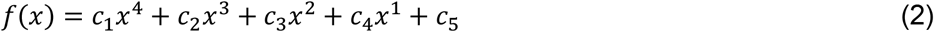

where *c*_*1*_, *c*_*2*_, …, *c*_*5*_ are the coefficients of the 4^th^ polynomial function *f(x)*. This polynomial fitting on the histogram (Supplementary Figure S3) provided overall information or a trend about all histogram frequency features.

### Spatial Analysis through Graph Neural Network

In this study, we developed a clustering-based spatial analysis framework using a graph deep learning model to extract key features of visual patterns from whole-slide imaging (WSI) of renal biopsies.^20,21^ In order to incorporate spatial information over the clustered image patterns on the biopsy sample, we spatially encoded clustered visual patterns on the original biopsy images with colors. The cluster indices obtained from the K-means clustering algorithm can be used to find the original location of each image tile on the biopsy sample. These spatially color-encoded visual patterns can be considered as nodes for the spatial analysis. In this study, to obtain the spatial neighborhood relationship between clustered image patterns on biopsy samples, we used a state-of-the-art graph neural network model called the DGCNN model.^41^

The DGCNN consists of four graph convolutional layers, a sort pooling layer, two 1-dimensional convolutional layers, a max pooling layer, and a fully connected layer. The representation of an entire graph can be obtained by summing the representation vectors of all nodes in the graph with a DGCNN algorithm (Figure 2).

**Figure 2.**
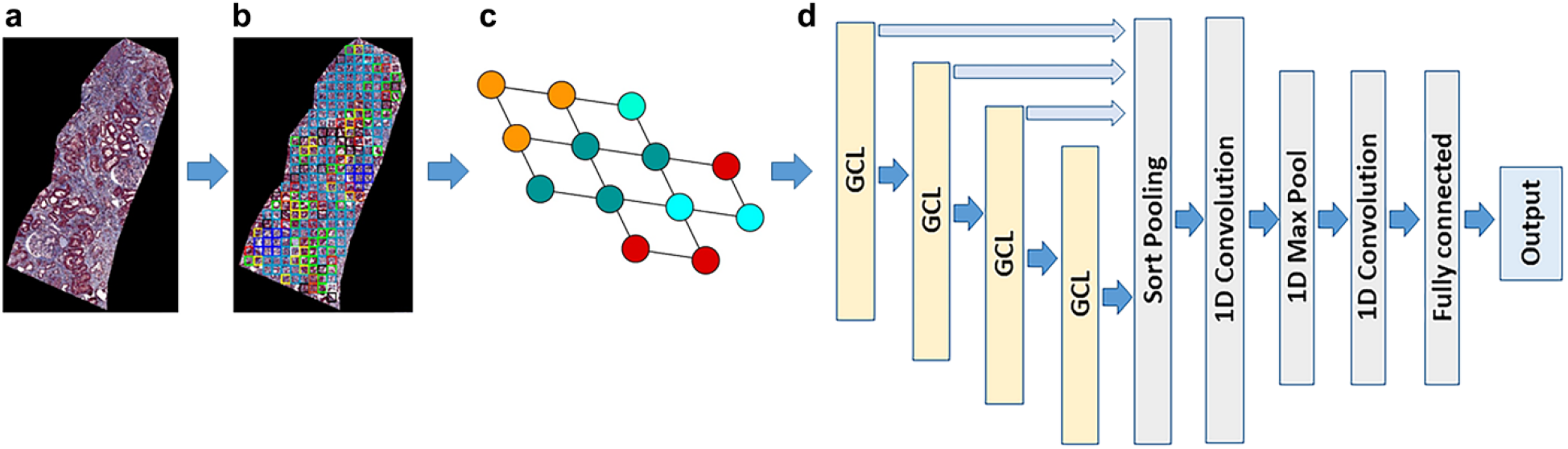
DCGNN architecture. (a) biopsy core image. (b) clustered color map. (c) graph representation of the clustered color map. (d) DCGNN architecture. The DCGNN model consisted of four graph convolutional layers, a sort pooling layer, two 1-dimensional convolutional layers, a max pooling layer, and a fully connected layer.

The graph representation as an input of the DGCNN was generated from the spatial location of visual patches using its adjacency and node features matrices. The node features were obtained using MDS, which calculates the dissimilarity between clustered visual pattern groups with the Euclidean distances to show how near clustered visual pattern groups are to each other.

The DCGNN outputs class prediction for each biopsy core image. Since each patient has multiple biopsy cores, we invented a normalized aw-score for the patient-level prediction which incorporates the normalized size of each core biopsy sample into its corresponding class prediction (1 or −1), then summed these aw-scores to get the patient-level outcome by Eq. (3).

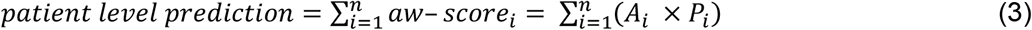

where *P* is an output of the predicted class either 1 or −1 from GDCNN for each biopsy core. *n* is the number of biopsy cores in each patient. *A* is the normalized area of each biopsy core. *aw-score* is defined as *A* × *P* for each core. We performed a 3-fold cross validation, stratified by patients (Tables 2 and 3), on DGCNN with listed area and prediction of DGCNN for each core as well as aw-scores for all patients obtained from the test sets of a 3-fold cross validation. For the spatial analysis with GNN, we used a python library for machine learning on graph-structured data, called StellarGraph based on TensorFlow and Keras API for the graph analysis.

### Random forest classifier with various groups of features

We obtained various groups of features: frequency or occurrence of each visual pattern group, polynomial fit coefficients from a histogram representation of clusters for each patient, spatial neighborhood relationship between clustered visual patterns, and clinical features such as age, race, and diagnosis. In this study, we used a random forest classifier with all features and divided the patients into two groups based on kidney function at the biopsy and one-year prediction, respectively. For the prediction at the biopsy, the patients were dichotomized into low and high eGFR groups with a threshold at eGFR 60: eGFR ≥ 60 (n=36) and eGFR < 60 (n=21). For the prediction whether eGFR is decreased or increased in one year, the patients were dichotomized into two groups based on the eGFR slope: eGFR slope ≥ 0 (n=30) and eGFR slope < 0 (n=27). The eGFR slope is defined as Eq. (4).

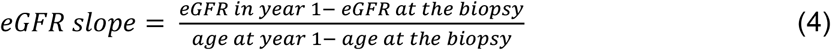

where “age at year 1” is the age in days approximately 1 year after the biopsy. A receiver operating characteristic (ROC) curve analysis was performed to illustrate the diagnostic ability of the binary classifier system. To evaluate the performance of our model, we estimated the area under the ROC curve (AUC) and its 95% confidence interval.^42-44^ The clustering analysis was performed using the algorithms “kmeans” and “silhouette” functions in MATLAB (R2020a, The MathWorks, Inc.). The ROC and AUC were computed using R (R Foundation for Statistical Computing, Vienna, Austria).

## RESULTS

### Unsupervised machine learning to cluster image patterns over the biopsy core images

In order to cluster image patterns on image patches, we extracted features from each image patch for the clustering using pre-trained DeepLab V3+ with ResNet-18 model.^38, 45^ Then, all 172 biopsy cores on images were tiled into 107,471 patches. Then, those patches were clustered through K-means clustering to group similar image patterns together (Figure 3a). K-means clustering algorithms use similarity or distance measures to form groupings such that image patches in the same groups are more similar than those in other groups. The optimal number of data clusters (K = 9) was obtained using the algorithm Silhouette in MATLAB (Supplementary Figure S2).

**Figure 3.**
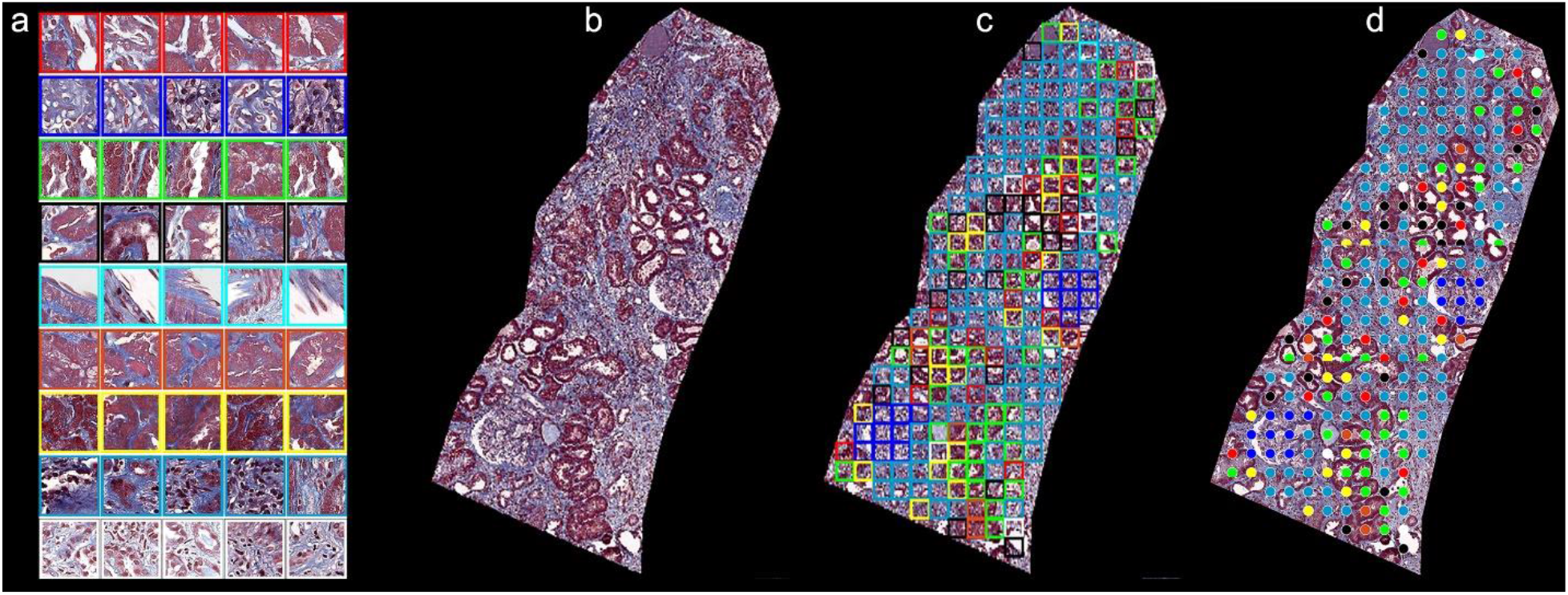
Clustered image patterns and examples. (a) clustered nine visual pattern groups with representative image patches; (b) an example of biopsy sample; (c) its cluster color-coded map; and (d) each color-coded patch can be considered a node for the spatial analysis.

Cluster indices, centroid locations, and distances from each point to every centroid were computed for the analysis. Figure 3 shows (a) 9 clustered visual pattern groups with representative image patches, (b) an example of cortexes, c) its color-coded cluster map, and (d) each color-coded patch can be considered a node for the spatial analysis. In this study, we developed a clustering-based deep learning methodology to find previously unknown visual patterns as well as spatial neighboring relationships between clustered kidney structure patterns that could predict patient outcomes. Table 1 summarized a detailed description for each visual pattern group and its importance for both predicting eGFR at the biopsy and in one year, respectively. The important visual patterns were established by using the Gini index, which calculates the amount of probability of a specific feature that is classified incorrectly when selected randomly.

**Table 1.** Description of 9 representative visual patterns

| Visual patterns | Color coded | Corresponding kidney structures | Rank (at the biopsy) | Rank (in one year) |
| --- | --- | --- | --- | --- |
| #1 | Red | Normal TI | 9 | 7 |
| #2 | Blue | Open glomerulus including normal and inflamed but not GS | 1 | 6 |
| #3 | Green | Normal TI – more white space or cells | 7 | 5 |
| #4 | Black | Normal TI, some interstitial expansion | 4 | 3 |
| #5 | Cyan | GS, IF, Arterioles including white space | 2 | 9 |
| #6 | Orange | Normal TI | 8 | 2 |
| #7 | Yellow | Normal and nearly normal TI with more interstitial area | 5 | 4 |
| #8 | Dark blue | Mostly interstitial expansion and tubular atrophy and some cellularity | 3 | 1 |
| #9 | White | Interstitial expansion | 6 | 8 |

TI: Tubulointerstitial; GS: Glomerulosclerosis; IF: Interstitial Fibrosis

### Graph neural network to incorporate the spatial information over the clustered image patterns on the biopsy core images

A graph for the GNN model is a data structure consisting of two components such as nodes and edges, which is used to analyze the pair-wise relationship between objects and entities. In this study, the graph representation as an input of the DGCNN was created with the nodes, which is the clustered visual patterns spatial located on the biopsy images.

In each case, there are multiple biopsy cores and the DCGNN outputs class prediction for each core and the patient-level prediction was obtained with a normalized area-weighted score, aw-score, by incorporating the normalized size of each core biopsy sample in a case into its corresponding patient-level prediction. We performed a 3-fold cross validation, stratified by patients, on DGCNN and Tables 2 and 3 listed normalized area and prediction of DGCNN for each core and aw-score for each case. These scores were obtained from the test sets from 3 different models (3-fold cross validation).

**Table 2.** aw-scores from the DGCNN for the prediction at the biopsy

| 3-fold groups | Case ID | # of cores | Normalized areas & core-level prediction (1 or -1) | aw-scores |
| --- | --- | --- | --- | --- |
| 1<sup>st</sup> group | 1 | 1 | 1 (1) | 1 |
|  | 2 | 3 | 0.10 (1), 0.28 (1), 0.16 (1) | 0.54 |
|  | 3 | 3 | 0.44(1), 0.56 (1), 0.40 (1) | 1.40 |
|  | 4 | 2 | 0.17(1), 0.20 (1) | 0.37 |
|  | 5 | 2 | 0.34 (1), 0.23 (1) | 0.57 |
|  | 6 | 5 | 0.09 (1), 0.17 (1), 0.16 (1), 0.02 (1), 0.04 (1) | 0.47 |
|  | 7 | 3 | 0.27 (1), 0.19 (1), 0.19 (1) | 0.66 |
|  | 8 | 3 | 0.24 (1), 0.40 (1), 0.38 (1) | 1.02 |
|  | 9 | 2 | 0.25 (-1), 0.59 (-1) | -0.84 |
|  | 10 | 2 | 0.79 (1), 0.28 (1) | 1.06 |
|  | 11 | 3 | 0.37 (-1), 0.57 (-1), 0.44 (-1) | -1.38 |
|  | 12 | 3 | 0.31 (1), 0.37 (1), 0.63 (1) | 1.30 |
|  | 13 | 1 | 0.54 (1) | 0.54 |
|  | 14 | 2 | 0.36 (1), 0.84 (-1) | -0.48 |
|  | 15 | 2 | 0.78 (1), 0.78 (1) | 1.56 |
|  | 16 | 3 | 0.56 (1), 0.67 (1), 0.50 (1) | 1.74 |
|  | 17 | 4 | 0.38 (1), 0.10 (1), 0.23 (1), 0.28 (1) | 0.99 |
|  | 18 | 2 | 0.44 (1), 0.41 (1) | 0.85 |
|  | 19 | 2 | 0.25 (1), 0.12 (1) | 0.38 |
| 2<sup>nd</sup> group | 20 | 4 | 0.28 (1), 0.04 (-1), 0.19 (1), 0.08 (1) | 0.51 |
|  | 21 | 2 | 0.59 (1), 0.74 (1) | 1.33 |
|  | 22 | 3 | 0.30 (-1), 0.08 (-1), 0.13 (-1) | -0.51 |
|  | 23 | 4 | 0.51 (-1), 0.18(-1), 0.39 (-1), 0.11 (-1) | -1.19 |
|  | 24 | 3 | 0.42 (1), 0.01 (-1), 0.47 (-1) | -0.06 |
|  | 25 | 4 | 0.17 (1), 0.08 (1), 0.05 (1), 0.08 (1) | 0.38 |
|  | 26 | 3 | 0.27 (-1), 0.07 (1), 0.36 (-1) | -0.55 |
|  | 27 | 1 | 0.13 (1) | 0.13 |
|  | 28 | 2 | 0.14 (1), 0.21 (1) | 0.35 |
|  | 29 | 3 | 0.08 (1), 0.04 (1), 0.22 (1) | 0.35 |
|  | 30 | 3 | 0.37 (1), 0.25 (1), 0.19 (1) | 0.81 |
|  | 31 | 2 | 0.37 (1), 0.27 (1) | 0.64 |
|  | 32 | 1 | 0.66 (-1) | -0.66 |
|  | 33 | 4 | 0.21 (1), 0.33 (1), 0.24 (1), 0.24 (1) | 1.03 |
|  | 34 | 2 | 0.85 (1), 0.96 (1) | 1.81 |
|  | 35 | 7 | 0.07 (-1), 0.02 (1), 0.01 (1), 0.01 (1), 0.07 (1), 0.01 (1), 0.01 (1) | 0.07 |
|  | 36 | 3 | 0.54 (-1), 0.40 (1), 0.52 (1) | 0.39 |
|  | 37 | 3 | 0.40 (-1), 0.14 (-1), 0.13 (-1) | -0.66 |
|  | 38 | 3 | 0.37 (-1), 0.20 (-1), 0.37 (-1) | -0.94 |
| 3<sup>rd</sup> group | 39 | 4 | 0.28 (-1), 0.23 (-1), 0.34 (-1), 0.33 (-1) | -1.18 |
|  | 40 | 2 | 0.26 (-1), 0.31 (-1) | -0.57 |
|  | 41 | 3 | 0.58 (-1), 0.41 (-1), 0.36 (-1) | -1.34 |
|  | 42 | 2 | 0.38 (-1), 0.30 (1) | -0.07 |
|  | 43 | 1 | 0.19 (1) | 0.19 |
|  | 44 | 4 | 0.30 (-1), 0.23 (-1), 0.32 (-1), 0.27 (-1) | -1.12 |
|  | 45 | 4 | 0.16 (1), 0.12 (1), 0.14 (1), 0.08 (1) | 0.49 |
|  | 46 | 5 | 0.53 (1), 0.26 (1), 0.06 (1), 0.54 (1), 0.47 (1) | 1.87 |
|  | 47 | 3 | 0.28 (1), 0.23 (1), 0.24 (1) | 0.75 |
|  | 48 | 1 | 0.58 (-1) | -0.58 |
|  | 49 | 2 | 0.46 (1), 0.52 (1) | 0.99 |
|  | 50 | 6 | 0.48 (-1), 0.32 (1), 0.23 (-1), 0.21 (-1), 0.23 (-1), 0.20 (1) | -0.64 |
|  | 51 | 3 | 0.42 (1), 0.05 (1), 0.36 (1) | 0.83 |
|  | 52 | 4 | 0.47 (-1), 0.16 (-1), 0.05 (-1), 0.46 (-1) | -1.14 |
|  | 53 | 5 | 0.43 (-1), 0.07 (1), 0.43 (1), 0.03 (1), 0.30 (-1) | -0.20 |
|  | 54 | 3 | 0.59 (-1), 0.34 (-1), 0.47 (-1) | -1.40 |
|  | 55 | 4 | 0.29 (-1), 0.32 (-1), 0.48 (-1), 0.42 (-1) | -1.50 |
|  | 56 | 4 | 0.17 (-1), 0.40 (-1), 0.04 (-1), 0.28 (-1) | -0.90 |
|  | 57 | 7 | 0.38 (-1), 0.18 (-1), 0.25 (-1), 0.28 (-1), 0.08 (-1), 0.01 (1), 0.04 (-1) | -1.21 |

**Table 3.** aw-scores from the DGCNN for the prediction in one year

| 3-fold groups | Case ID | # of cores | Normalized areas & core-level prediction (1 or -1) | aw-scores |
| --- | --- | --- | --- | --- |
| 1<sup>st</sup> group | 1 | 1 | 1 (1) | 1 |
|  | 2 | 3 | 0.10 (1), 0.28 (1), 0.16 (-1) | 0.22 |
|  | 3 | 3 | 0.44 (1), 0.56 (1), 0.40 (1) | 1.40 |
|  | 4 | 2 | 0.17 (1), 0.20 (1) | 0.37 |
|  | 5 | 2 | 0.34 (1), 0.23 (1) | 0.11 |
|  | 6 | 5 | 0.09 (1), 0.17 (1), 0.16 (1), 0.02 (1), 0.04 (1) | 0.47 |
|  | 7 | 3 | 0.27 (1), 0.19 (1), 0.19 (1) | 0.66 |
|  | 8 | 3 | 0.24 (1), 0.39 (1), 0.38 (1) | 1.02 |
|  | 9 | 2 | 0.25 (-1), 0.59 (-1) | -0.84 |
|  | 10 | 2 | 0.79 (-1), 0.28 (-1) | -1.06 |
|  | 11 | 3 | 0.37 (-1), 0.57 (-1), 0.44 (-1) | -1.38 |
|  | 12 | 3 | 0.31 (1), 0.37 (1), 0.63 (1) | 1.30 |
|  | 13 | 1 | 0.54 (-1) | -0.54 |
|  | 14 | 2 | 0.36 (-1), 0.84 (-1) | -1.19 |
|  | 15 | 2 | 0.78 (1), 0.79 (-1) | -0.01 |
|  | 16 | 3 | 0.56 (-1), 0.67 (-1), 0.50 (1) | -0.73 |
|  | 17 | 4 | 0.38 (-1), 0.10 (1), 0.23 (1), 0.28 (1) | 0.22 |
|  | 18 | 2 | 0.44 (-1), 0.41 (1) | -0.03 |
|  | 19 | 2 | 0.25 (1), 0.12 (-1) | 0.13 |
| 2<sup>nd</sup> group | 20 | 4 | 0.28 (-1), 0.04 (-1), 0.19 (-1), 0.08 (1) | -0.43 |
|  | 21 | 2 | 0.59 (1), 0.74 (1) | 1.33 |
|  | 22 | 3 | 0.30 (-1), 0.08 (-1), 0.13 (-1) | -0.51 |
|  | 23 | 4 | 0.51 (-1), 0.18 (-1), 0.39 (-1), 0.11 (-1) | -1.19 |
|  | 24 | 3 | 0.42 (-1), 0.01 (-1), 0.47 (-1) | -0.90 |
|  | 25 | 4 | 0.17 (-1), 0.08 (-1), 0.05 (1), 0.08 (1) | -0.12 |
|  | 26 | 3 | 0.27 (-1), 0.07 (-1), 0.36 (-1) | -0.70 |
|  | 27 | 1 | 0.13 (-1) | -0.13 |
|  | 28 | 2 | 0.14 (1), 0.21 (1) | 0.35 |
|  | 29 | 3 | 0.08 (-1), 0.04 (-1), 0.22 (-1) | -0.35 |
|  | 30 | 3 | 0.37 (1), 0.25 (1), 0.19 (1) | 0.81 |
|  | 31 | 2 | 0.37 (-1), 0.27 (-1) | -0.64 |
|  | 32 | 1 | 0.66 (-1) | -0.66 |
|  | 33 | 4 | 0.21 (-1), 0.33 (1), 0.24 (-1), 0.24 (1) | 0.11 |
|  | 34 | 2 | 0.85 (1), 0.96 (1) | 1.81 |
|  | 35 | 7 | 0.07 (-1), 0.02 (1), 0.01 (1), 0.01 (1), 0.07 (-1), 0.01 (-1), 0.01 (1) | -0.12 |
|  | 36 | 3 | 0.54 (-1), 0.40 (1), 0.52 (1) | 0.39 |
|  | 37 | 3 | 0.40 (-1), 0.14 (-1), 0.13 (-1) | -0.66 |
|  | 38 | 3 | 0.37 (-1), 0.20 (-1), 0.37 (-1) | -0.94 |
| 3<sup>rd</sup> group | 39 | 4 | 0.28 (-1), 0.23 (-1), 0.34 (-1), 0.33 (-1) | -1.18 |
|  | 40 | 2 | 0.26 (-1), 0.31 (-1) | -0.57 |
|  | 41 | 3 | 0.58 (-1), 0.41 (-1), 0.36 (-1) | -1.34 |
|  | 42 | 2 | 0.38 (-1), 0.30 (1) | -0.07 |
|  | 43 | 1 | 0.19 (-1) | -0.19 |
|  | 44 | 4 | 0.30 (-1), 0.23 (-1), 0.32 (-1), 0.27 (-1) | -1.12 |
|  | 45 | 4 | 0.16 (1), 0.12 (-1), 0.14 (1), 0.08 (1) | 0.26 |
|  | 46 | 5 | 0.53 (-1), 0.26 (1), 0.06 (1), 0.54 (-1), 0.47 (-1) | -1.23 |
|  | 47 | 3 | 0.28 (1), 0.23 (1), 0.24 (1) | 0.75 |
|  | 48 | 1 | 0.58 (-1) | -0.58 |
|  | 49 | 2 | 0.47 (1), 0.52 (1) | 0.99 |
|  | 50 | 6 | 0.48 (-1), 0.32 (-1), 0.23 (-1), 0.21 (-1), 0.23 (-1), 0.20 (-1) | -1.67 |
|  | 51 | 3 | 0.42 (-1), 0.05 (1), 0.36 (1) | -0.02 |
|  | 52 | 4 | 0.48 (-1), 0.16 (-1), 0.05 (-1), 0.46 (-1) | -1.14 |
|  | 53 | 5 | 0.43 (-1), 0.07 (-1), 0.43 (-1), 0.03 (-1), 0.30 (-1) | -1.25 |
|  | 54 | 3 | 0.59 (-1), 0.34 (-1), 0.47 (-1) | -1.40 |
|  | 55 | 4 | 0.29 (-1), 0.32 (-1), 0.48 (-1), 0.42 (-1) | -1.50 |
|  | 56 | 4 | 0.17 (-1), 0.40 (-1), 0.04 (-1), 0.28 (-1), 0.39 (-1) | -0.90 |
|  | 57 | 7 | 0.38 (-1), 0.18 (-1), 0.25 (-1), 0.28 (-1), 0.08 (-1), 0.01 (1), 0.04 (-1) | -1.21 |

### Machine learning classifier with cluster frequency features, polynomial fitting features, clinical features, and aw-scores from GNN

A histogram representation of clusters for each patient was created to describe the distribution of each type of cluster at the patient level. This cluster frequency information from the histogram gives us the cluster frequency features for each patient. Investigating these visual patterns for each case can give us information to find previously unknown features that predict patient outcomes (Table 1). Some visual pattern groups match well with distinct microscopic kidney structures: Visual pattern group #2 (blue nodes) is the glomerular structure, which is the most important visual pattern for predicting kidney function at the biopsy and visual pattern group #5 is arterioles with some white space. Visual pattern group #7 does not exactly match with distinctive microscopic kidney structures but contains both normal and near normal tubulointerstitial (TI) and some interstitial areas, which are the most important visual patterns for predicting kidney function in one year. Figure 4 shows two examples of biopsy samples; one (right) has complex heterogeneous visual patterns and one (left) has relatively few and distinctive visual patterns. Through our CluSA framework, we can assess complex heterogeneous visual patterns of biopsy samples not only through their quantities in CKD patient tissue but also through their spatial configuration in the tissue.

**Figure 4.**
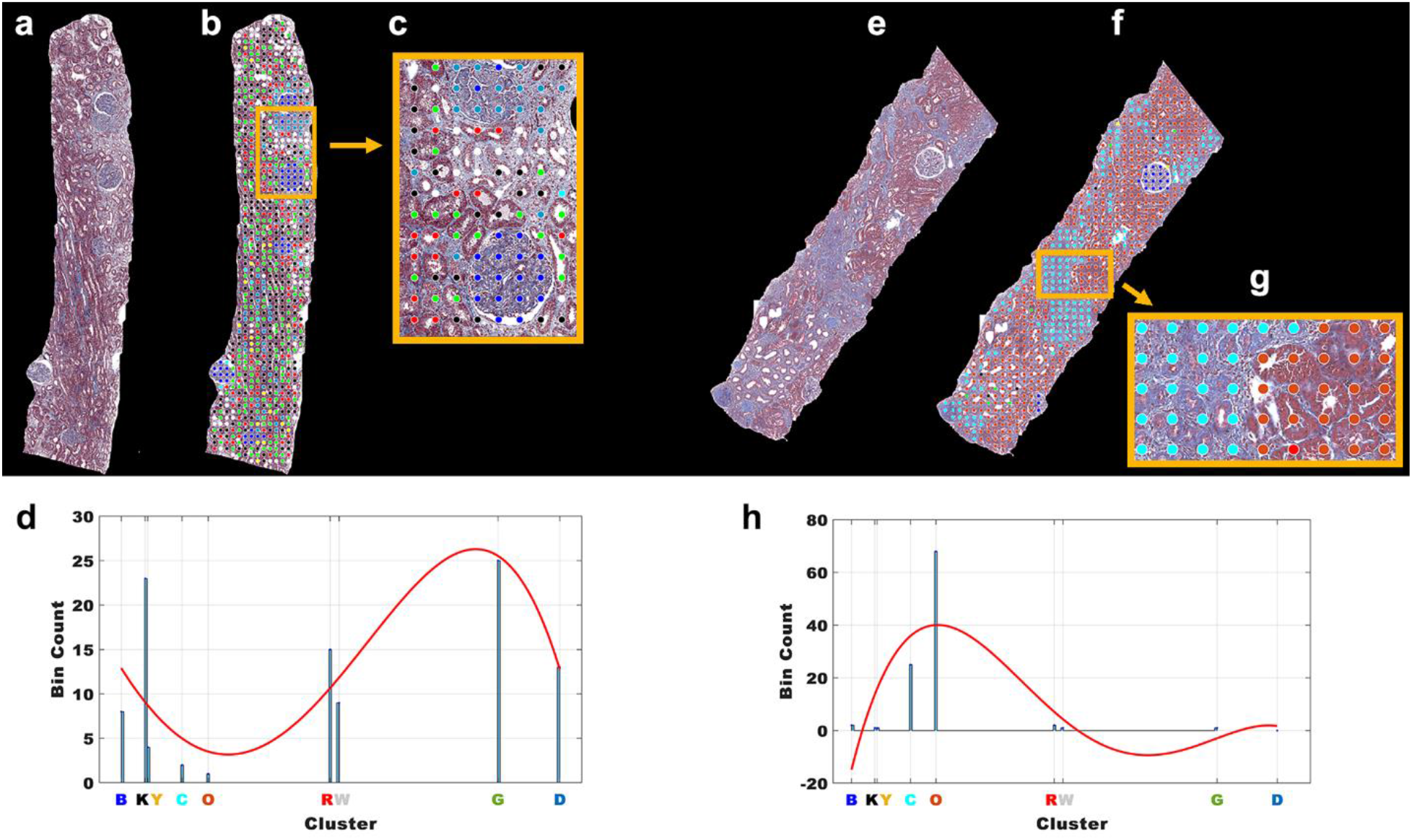
Two examples of biopsy samples. (a) and (e) two examples of biopsy samples; (b) and (f) color-coded nodes from clustered visual pattern groups; (c) and (g) zoom images; (d) and (h) histogram representation of clusters with 4^th^ polynomial fitting curves for both cases, respectively.

In addition to the individual frequency or occurrence of visual patterns on the histogram, the polynomial fitting on the histogram provided overall information about all histogram cluster frequency features (Figure 4 (d, h) and Supplementary Figure S3). In this study, we used a machine learning classifier to incorporate all features such as histogram frequency features, polynomial coefficient features, aw-scores from DGCNN, and clinical features such as age, race, gender, and diagnosis to predict patient outcomes. The detailed clinical features including patient diagnosis and demographics are shown in Supplementary Table S1. We used a random forest model as a classifier to calculate AUC and predict association of visual patterns and features with clinical patient outcomes such as eGFR. Tables 4 and 5 show the ranking of the important features for the dichotomized level of kidney function at the biopsy and in one year, respectively. We used additional clinical features, eGFR and UPC at the biopsy, for the prediction of eGFR changes in one year. These important features were computed by using the Gini index, also known as Gini impurity, which calculates the amount of probability of a specific feature that is classified incorrectly when selected randomly.

**Table 4.** Ranking of the important features for the level of kidney function at the biopsy.

| Features | Description<br>(* top 7 features) | Gini index<br>(importance) | Rank | Overall Rank |
| --- | --- | --- | --- | --- |
| Spatial feature | aw-scores* | 3.04 | 1 | 1 |
| Frequency<br>(visual dictionary) | f1 (red) | 0.77 | 9 | 17 |
|  | f2 (blue)* | 2.31 | 1 | 3 |
|  | f3 (green) | 0.97 | 7 | 14 |
|  | f4 (black) | 1.35 | 4 | 8 |
|  | f5 (cyan)* | 1.77 | 2 | 5 |
|  | f6 (orange) | 0.92 | 8 | 15 |
|  | f7 (yellow) | 1.14 | 5 | 11 |
|  | f8 (dark blue)* | 1.38 | 3 | 7 |
|  | f9 (white) | 1.06 | 6 | 13 |
| Polynomial coefficient | $c_1$* | 2.15 | 1 | 4 |
| | $c_2$ | 1.19 | 2 | 9 |
| | $c_3$ | 1.11 | 4 | 12 |
| | $c_4$ | 1.14 | 3 | 10 |
| | $c_5$ | 0.84 | 5 | 16 |
| Clinical features | Age* | 2.66 | 1 | 2 |
|  | Gender | 0.45 | 4 | 19 |
|  | Race | 0.48 | 3 | 18 |
|  | Diagnosis* | 1.46 | 2 | 6 |

$c_n$ : polynomial coefficients in Eq. [1];

**Table 5.** Ranking of the important features for the prediction of eGFR changes in one year.

| Features | Description<br>(* top 7 features) | Gini index<br>(importance) | Rank | Overall Rank |
| --- | --- | --- | --- | --- |
| Spatial feature | aw-scores* | 3.30 | 1 | 1 |
| Frequency<br>(visual patterns) | f1 (red) | 0.92 | 7 | 16 |
|  | f2 (blue) | 1.31 | 6 | 10 |
|  | f3 (green) | 1.36 | 5 | 9 |
|  | f4 (black)* | 1.45 | 3 | 7 |
|  | f5 (cyan) | 0.84 | 9 | 18 |
|  | f6 (orange) | 1.79 | 2 | 4 |
|  | f7 (yellow) | 1.42 | 4 | 8 |
|  | f8 (dark blue)* | 2.16 | 1 | 2 |
|  | f9 (white) | 0.86 | 8 | 17 |
| Polynomial coefficient | $c_1$ | 1.30 | 3 | 11 |
| | $c_2$* | 1.64 | 2 | 5 |
| | $c_3$ | 1.21 | 4 | 12 |
| | $c_4$* | 2.00 | 1 | 3 |
| | $c_5$ | 1.17 | 5 | 14 |
| Clinical features | Age* | 1.59 | 1 | 6 |
|  | Gender | 0.17 | 6 | 21 |
|  | Race | 0.36 | 5 | 20 |
|  | Diagnosis | 0.82 | 4 | 19 |
|  | eGFR | 1.13 | 3 | 15 |
|  | UPC | 1.19 | 2 | 13 |

$c_n$ : polynomial coefficients in Eq. [1]; **UPC**: Urine Protein Creatinine ratio;

We selected the top 7 most important features based on the importance index rank (Tables 4 and 5): one spatial feature, three frequency features (f2, f5, and f8), one polynomial feature (c1), and two clinical features (age and diagnosis) for predicting eGFR at the biopsy; and one spatial feature, three frequency features (f4, f6, and f8), two polynomial feature (c2 and c4), and one clinical feature (age) for predicting eGFR changes in 1 year. Selecting the top 7 features ensured that all four categories of feature types were included in our analysis. For predicting eGFR at the biopsy, the error from the random forest was 0.05, the sensitivity was 0.97, and specificity was 0.90. The ROC for the top 7 features is illustrated in Figure 5(a). The AUC was 0.96 and the 95% confidence interval was 0.89 – 1.0. The accuracy was 0.95.

**Figure 5.**
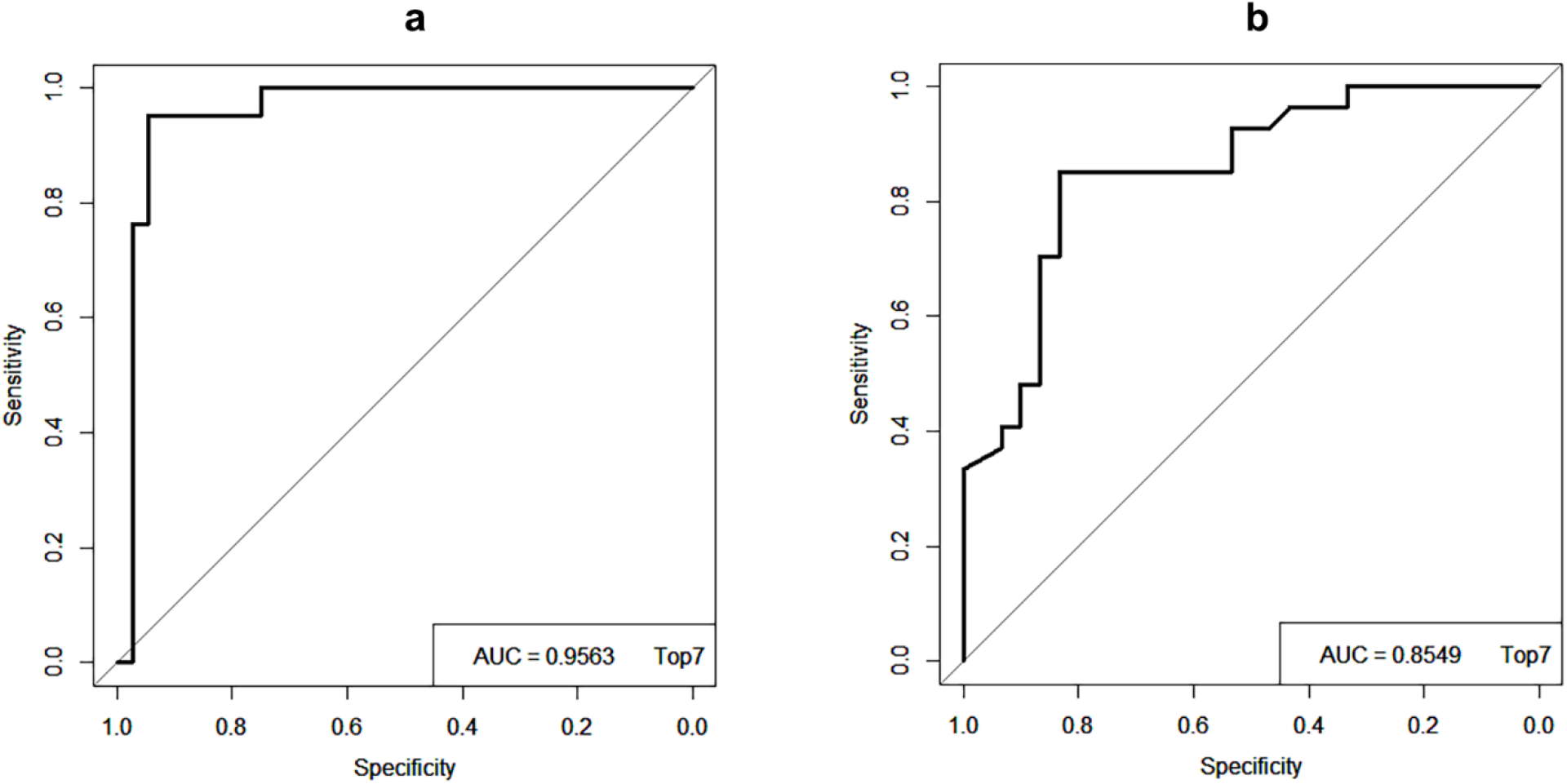
ROC curves and AUC values. ROC curves for the prediction of the level of kidney function, (a) at the biopsy and, (b) in the future. Top7 represents the top 7 features selected based on the importance rank. The x-axis is the true negative rate (TNR) or specificity and the y-axis is the true positive rate (TPR) or sensitivity.

For predicting whether eGFR is increased or decreased in one year, the error from the random forest was 0.16; sensitivity, 0.83; specificity, 0.85; and accuracy, 0.84. The ROC for the top 7 features is illustrated in Figure 5 (b). The area under the ROC curve (AUC) was 0.85 and the 95% confidence interval was 0.76 – 0.95. The accuracies were calculated by using Eq. (1) for this model,

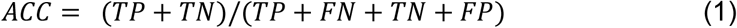

where, TP, FP, TN, and FN represent true-positive, false-positive, true-negative, and false-negative predictions, respectively. The detailed results of confusion matrix, AUC, 95% Confidence Interval (CI), and accuracy are shown in Supplementary Table S2 and S3, respectively. Based on the results, the spatial feature of neighborhood information between clustered visual patterns from the graph neural network was the most important feature associated with the prediction of the kidney function at the biopsy as well as in one year.

## DISCUSSION

In histopathology image analysis, artificial intelligence and machine learning methods have been used in computer-aided studies to solve diagnostic decision-making problems, and most of the machine learning methods applied to histopathology slides have relied on fully supervised learning and pixel-level expert annotations to extract features or train a model.^11,13,29,30^ However, supervised machine learning presents some major challenges. First, supervised learning requires significant labeling efforts, which is a very time-consuming task that is often impractical in histopathology images due to their large image size with high resolution. Also, labeling microstructures or regions of interests (ROIs) on histopathology images requires the domain knowledge of the microstructures of ROIs as well. Further, the model’s results depend on the quality of labels in the training set, which could involve human error in manual labeling. On the other hand, unsupervised learning does not require labeled data and the model learns from raw data without any prior knowledge. In addition, it discovers previously unknown patterns from the data. However, unsupervised learning has some disadvantages as well. One of the typical disadvantages is that it does not consider spatial relationships in the data.

In order to build highly intelligent and efficient machine learning algorithms, we developed a computational framework that uses unsupervised learning to overcome the burden of manual labeling and supervised machine learning to incorporate a spatial relationship between visual patterns. The best way to make progress on this is through unsupervised machine learning using a clustering algorithm, which does not require labeled data, and find the original location of each clustered patch on the biopsy sample image. Also, clustering-based analysis has no specific sample size limitation. As shown in Figure 3, each clustered color-coded patch can be considered a node, and a graph representation was obtained from these nodes for the spatial analysis. In this study, we identified the most important features among normalized aw-score feature that contains spatial information between neighboring image patterns, cluster frequency features that represent a quantitative amount of each clustered image pattern or node within a case, polynomial fit coefficient features that provide overall information or a trend about all cluster frequency features, and clinical features that include age, race, and diagnosis. This is done by computing the Gini index or Gini impurity. The most important feature was the normalized aw-score feature obtained from the graph deep learning model for both predictions at the biopsy and in one year. This shows that the spatial pattern of neighboring image patterns or fibrosis, which is a characteristic feature of all forms of CKD,^31^ could be an important factor to be considered for the level of kidney function in CKD. To our knowledge, CluSA is the first study in which unsupervised machine learning has been used to cluster morphologic visual patterns and assess the spatial neighborhood relationship between clustered visual patterns to predict the kidney function in CKD.

Our retrospective study has a couple of limitations. First, in this study we fixed the patch size at 256 × 256 pixels based on the requirement of the input image size of the pretrained deep learning model. However, other image sizes with rescaling or with some degree of overlap between adjacent patches could be investigated in the future study. Secondly, although k-means clustering is one of the popular unsupervised learning methods to cluster unlabeled data into k clusters, identifying an optimal number of clusters in a dataset is a fundamental issue and there is no definitive answer as to the true number of clusters. To determine the optimal number of clusters in k-means clustering, we used one of the most popular algorithms, silhouette, which measures the quality of a clustering. Its value indicates a measure of how similar an object is to its own cluster compared to other clusters. However, the effects of the number of clusters on the clustering performance should be explored in a future study with different patch size. Similarly, future studies that provide more systematic examination of the effects of stain color normalization and various staining such as H&E and PAS are needed.

Previously, we have shown that unsupervised machine-learned clustering features are potential surrogates of predicting eGFR and can be used as tools for prognosis as well as for objective assessment of the level of kidney function in CKD^45^. In the present study, our results demonstrate that the addition of spatial information improves the model’s performance by 2.4% and 5.1% of AUC at the biopsy and one-year prediction, respectively, compared to the previous study. Furthermore, we identified that aw-score, consisting of results from the GNN model, is the most important feature for predicting patient outcomes. The clustering of visual patterns enables pathologists to inspect these key image segments for clinically significant data. In contrast to traditional deep learning approaches, in which an algorithm learns from data labeled by a pathologist with known pathologic features for classifying disease, this unsupervised approach via CluSA automatically identifies the most optimal discriminative features, some of which may be potentially new, for understanding and prognosticating disease.

It is important that our framework can find important visual patterns of the kidney tissue corresponding to the patient outcomes without human input and can predict future diagnosis. Although further study is required for complex disease analysis, our computational CluSA framework will potentially have benefits from having a higher speed of execution, accuracy, and incorporating spatial information while minimizing the need for time-consuming, impractical expert annotations.

## CONCLUSION

In this study, we showed that the identifying morphological characteristics from clustering and the spatial relationship between them can not only remove the burden of obtaining manual-labeled training datasets, but also provide interpretability in the form of spatial visualizations of predictive features. The results from our study also indicate that the spatial relationship between visual patterns obtained from unsupervised machine learning is the most important feature that can predict outcomes. Our objective computational CluSA framework will be useful for discriminating levels of kidney function as well as other disease in digital histopathology image analysis. Since clustering-based analysis has no sample size limitation, our CluSA framework confers real practical use with relatively small datasets and could help in decision making during follow-up.

## Data Availability

All data produced in the present study are available upon reasonable request to the authors

## ACKNOWLEDGEMENTS

We would like to thank all staffs at the J.B.H. lab for their contribution to this study. This study was supported by Department of Defense (DoD) grants W81XWH2210032 (J.L.) and W81XWH2010436 (J.B.H & A.R) as well as NCI Grant R37-CA214955. A.R. and E.W. was supported by CCSG Bioinformatics Shared Resource 5 P30 CA046592, a gift from Agilent technologies, a Research Scholar Grant from the American Cancer Society (RSG-16-005-01), and a Precision Health Investigator award from U-M Precision Health to A.R. along with L. Rozek and M. Sartor. J.L, E.W. and A.R. were partially supported by The University of Michigan (U-M) startup institutional research funds.

## AUTHOR CONTRIBUTIONS

Project conception and design were by J.L., J.B.H., and A.R. The data collection and preprocessing were performed by J.L., E.W., S.S., M.B., M.K., D.G., S.P., K.B., Z.B., C.G., S.M., K.P., J.S., Y.Y., J.L., X.Z., L.M., and J.B.H. The software programming, statistical and data analysis, and interpretation were performed by J.L. The manuscript was written by J.L. and all authors reviewed the manuscript.

## REFERENCES

1. Centers for Disease Control and Prevention. Chronic Kidney Disease Surveillance System website. https://nccd.cdc.gov/CKD Accessed June 8. 2020. In (vol 2020)

2. Romagnani P, Remuzzi G, Glassock R, et al. Chronic kidney disease. Nat Rev Dis Primers 2017; 3: 17088.

3. Gansevoort RT, Matsushita K, van der Velde M, et al. Lower estimated GFR and higher albuminuria are associated with adverse kidney outcomes. A collaborative meta-analysis of general and high-risk population cohorts. Kidney Int 2011; 80: 93–104.

4. Qaseem A, Hopkins RH, Jr., Sweet DE, et al. Screening, monitoring, and treatment of stage 1 to 3 chronic kidney disease: A clinical practice guideline from the American College of Physicians. Ann Intern Med 2013; 159: 835–847.

5. da Silva Selistre L, Rech DL, de Souza V, et al. Diagnostic Performance of Creatinine-Based Equations for Estimating Glomerular Filtration Rate in Adults 65 Years and Older. JAMA Intern Med 2019; 179: 796–804.

6. Tangri N, Stevens LA, Griffith J, et al. A predictive model for progression of chronic kidney disease to kidney failure. JAMA 2011; 305: 1553–1559.

7. Levey AS, Stevens LA, Schmid CH, et al. A new equation to estimate glomerular filtration rate. Ann Intern Med 2009; 150: 604–612.

8. Nath KA. Tubulointerstitial changes as a major determinant in the progression of renal damage. Am J Kidney Dis 1992; 20: 1–17.

9. Bhargava R, Madabhushi A. Emerging Themes in Image Informatics and Molecular Analysis for Digital Pathology. Annu Rev Biomed Eng 2016; 18: 387–412.

10. Coudray N, Ocampo PS, Sakellaropoulos T, et al. Classification and mutation prediction from non-small cell lung cancer histopathology images using deep learning. Nat Med 2018; 24: 1559–1567.

11. Bueno G, Fernandez-Carrobles MM, Gonzalez-Lopez L, et al. Glomerulosclerosis identification in whole slide images using semantic segmentation. Comput Methods Programs Biomed 2020; 184: 105273.

12. Kannan S, Morgan LA, Liang B, et al. Segmentation of Glomeruli Within Trichrome Images Using Deep Learning. Kidney Int Rep 2019; 4: 955–962.

13. Kolachalama VB, Singh P, Lin CQ, et al. Association of Pathological Fibrosis With Renal Survival Using Deep Neural Networks. Kidney Int Rep 2018; 3: 464–475.

14. Sudharshan PJ, Petitjean C, Spanhol F, et al. Multiple instance learning for histopathological breast cancer image classification. Expert Syst Appl 2019; 117: 103–111.

15. Vu T, Lai P, Raich R, et al. A Novel Attribute-Based Symmetric Multiple Instance Learning for Histopathological Image Analysis. Ieee T Med Imaging 2020; 39: 3125–3136.

16. Xu Y, Zhu JY, Chang EI, et al. Weakly supervised histopathology cancer image segmentation and classification. Med Image Anal 2014; 18: 591–604.

17. Kanavati F, Toyokawa G, Momosaki S, et al. Weakly-supervised learning for lung carcinoma classification using deep learning. Sci Rep 2020; 10: 9297.

18. van der Laak J, Litjens G, Ciompi F. Deep learning in histopathology: the path to the clinic. Nat Med 2021; 27: 775–784.

19. Schapiro D, Jackson HW, Raghuraman S, et al. histoCAT: analysis of cell phenotypes and interactions in multiplex image cytometry data. Nat Methods 2017; 14: 873–876.

20. Heindl A, Nawaz S, Yuan Y. Mapping spatial heterogeneity in the tumor microenvironment: a new era for digital pathology. Lab Invest 2015; 95: 377–384.

21. Zhang Z, Chen L, Zhong F, et al. Graph neural network approaches for drug-target interactions. Curr Opin Struct Biol 2022; 73: 102327.

22. Xuan P, Pan S, Zhang T, et al. Graph Convolutional Network and Convolutional Neural Network Based Method for Predicting lncRNA-Disease Associations. Cells 2019; 8.

23. Peng H, Li JX, He Y, et al. Large-Scale Hierarchical Text Classification with Recursively Regularized Deep Graph-CNN. Web Conference 2018: Proceedings of the World Wide Web Conference (www2018) 2018: 1063-1072.

24. Veličković P, Cucurull G, Casanova A, et al. Graph attention networks. 2017.

25. Lopez C, Tucker S, Salameh T, et al. An unsupervised machine learning method for discovering patient clusters based on genetic signatures. J Biomed Inform 2018; 85: 30–39.

26. Levey AS, Stevens LA. Estimating GFR Using the CKD Epidemiology Collaboration (CKD-EPI) Creatinine Equation: More Accurate GFR Estimates, Lower CKD Prevalence Estimates, and Better Risk Predictions. Am J Kidney Dis 2010; 55: 622–627.

27. Reinhard E, Ashikhmin N, Gooch B, et al. Color transfer between images. Ieee Comput Graph 2001; 21: 34–41.

28. Routray S, Ray AK, Mishra C. Analysis of Various Image Feature Extraction Methods against Noisy Image: SIFT, SURF and HOG. Proceedings of the 2017 Ieee Second International Conference on Electrical, Computer and Communication Technologies (Icecct) 2017.

29. Kumar G, Bhatia PK. A Detailed Review of Feature Extraction in Image Processing Systems. Int C Adv Comput Com 2014: 5-+.

30. Cheplygina V, de Bruijne M, Pluim JPW. Not-so-supervised: A survey of semi-supervised, multi-instance, and transfer learning in medical image analysis. Med Image Anal 2019; 54: 280–296.

31. Liu SP, Tian GH, Xu Y. A novel scene classification model combining ResNet based transfer learning and data augmentation with a filter. Neurocomputing 2019; 338: 191–206.

32. Morid MA, Borjali A, Del Fiol G. A scoping review of transfer learning research on medical image analysis using ImageNet. Comput Biol Med 2021; 128: 104115.

33. Shin HC, Roth HR, Gao M, et al. Deep Convolutional Neural Networks for Computer-Aided Detection: CNN Architectures, Dataset Characteristics and Transfer Learning. IEEE Trans Med Imaging 2016; 35: 1285–1298.

34. van Opbroek A, Ikram MA, Vernooij MW, et al. Transfer learning improves supervised image segmentation across imaging protocols. IEEE Trans Med Imaging 2015; 34: 1018–1030.

35. Christopher M, Belghith A, Bowd C, et al. Performance of Deep Learning Architectures and Transfer Learning for Detecting Glaucomatous Optic Neuropathy in Fundus Photographs. Sci Rep 2018; 8: 16685.

36. Pratt LY (ed). Discriminability-based transfer between neural networks. Proceedings of he Conference Name; Date Year of Conference; Conference Location|. Publisher|: Place Published|, Year Published|.

37. Chen LCE, Zhu YK, Papandreou G, et al. Encoder-Decoder with Atrous Separable Convolution for Semantic Image Segmentation. Lect Notes Comput Sc 2018; 11211: 833–851.

38. He KM, Zhang XY, Ren SQ, et al. Deep Residual Learning for Image Recognition. Proc Cvpr Ieee 2016: 770–778.

39. Russakovsky O, Deng J, Su H, et al. ImageNet Large Scale Visual Recognition Challenge. Int J Comput Vision 2015; 115: 211–252.

40. He K, Zhang X, Ren S, et al. (eds). Deep residual learning for image recognition. Proceedings of he Conference Name; Date Year of Conference; Conference Location|. Publisher|: Place Published|, Year Published|.

41. Zhang MH, Cui ZC, Neumann M, et al. An End-to-End Deep Learning Architecture for Graph Classification. Thirty-Second Aaai Conference on Artificial Intelligence / Thirtieth Innovative Applications of Artificial Intelligence Conference / Eighth Aaai Symposium on Educational Advances in Artificial Intelligence 2018: 4438-4445.

42. Bradley AP. The use of the area under the roc curve in the evaluation of machine learning algorithms. Pattern Recogn 1997; 30: 1145–1159.

43. Khatun MS, Shoombuatong W, Hasan MM, et al. Evolution of Sequence-based Bioinformatics Tools for Protein-protein Interaction Prediction. Curr Genomics 2020; 21: 454–463.

44. Khatun MS, Alam MA, Shoombuatong W, et al. Recent development of bioinformatics tools for microRNA target prediction. Curr Med Chem 2021.

45. Lee J, Warner E, Shaikhouni S, et al. Unsupervised machine learning for identifying important visual features through bag-of-words using histopathology data from chronic kidney disease. Sci Rep 2022; 12: 4832.

