## Supplementary for "CluSA: Clustering-based Spatial Analysis framework through Graph Neural Network for Chronic Kidney Disease Prediction using Histopathology Images"

### SUPPLIMENTARY MATERIAL

**Table S1**. Clinical data for 57 CKD patients[1].

| Patients | Gender | Race | Diagnosis | eGFR | UPC |
|---|---|---|---|---|---|
| 1 | Female | White/Caucasian | Lupus Class III | 96.87 | 2.27 |
| 2 | Female | White/Caucasian | Lupus Class V, MPGN | 90.93 | 1.46 |
| 3 | Female | White/Caucasian | Lupus Class V | 118.92 | 1.9 |
| 4 | Female | Black/African American | Lupus Class III, Lupus Class IV | 113.06 | 0.43 |
| 5 | Female | Asian/Asian American | Lupus Class III | 121.27 | 1.87 |
| 6 | Male | White/Caucasian | Minimal Change | 70.58 | 7.71 |
| 7 | Female | White/Caucasian | HSP | 122.12 | 0.48 |
| 8 | Male | White/Caucasian | FSGS, IgA Nephropathy | 47.31 | 2.2 |
| 9 | Female | American Indian/Alaskan Native | FSGS | 119.68 | 0.9 |
| 10 | Female | White/Caucasian | Diabetic nephropathy | 26.62 | 4.6 |
| 11 | Female | White/Caucasian | Minimal change, FSGS | 27.63 | 3.36 |
| 12 | Male | Black/African American | FSGS | 70.82 | 1.27 |
| 13 | Male | White/Caucasian | FSGS | 118.9 | 1.86 |
| 14 | Female | Black/African American | Lupus Class V, hypertensive nephropathy | 43.59 | 0.15 |
| 15 | Female | Black/African American | Lupus Class III, lupus Class V | 98.93 | 8.25 |
| 16 | Female | White/Caucasian | Lupus nephritis | 79.98 | 5.37 |
| 17 | Female | White/Caucasian | HSP | 160.93 | 1.72 |
| 18 | Female | White/Caucasian | IgA Nephropathy | 93.52 | 5.97 |
| 19 | Female | White/Caucasian | Lupus Class II, MPGN | 104.65 | 2.69 |
| 20 | Female | Black/African American | Lupus Class V | 83.35 | 2.48 |
| 21 | Female | White/Caucasian | Lupus Class III | 107.78 | 2.9 |
| 22 | Female | White/Caucasian | FSGS | 32.67 | 0.21 |
| 23 | Male | Asian/Asian American | IgA Nephropathy, Hypertensive nephropathy | 34.4 | 2.95 |
| 24 | Male | White/Caucasian | Glomerular disease | 22.96 | 2.23 |
| 25 | Female | White/Caucasian | Membranous Nephropathy | 105.13 | 3.9 |
| 26 | Male | White/Caucasian | Glomerular disease | 96.94 | 0.08 |
| 27 | Female | Black/African American | Lupus class IV | 95.905 | 1.65 |
| 28 | Male | White/Caucasian | FSGS, tubulointerstitial disease | 30.8 | 8.13 |
| 29 | Female | Multiracial | Lupus class III, Lupus class IV | 142.74 | 0.73 |
| 30 | Male | White/Caucasian | IgA Nephropathy | 131.41 | 3.41 |
| 31 | Female | Black/African American | Lupus Class V | 117.93 | 0 |
| 32 | Female | White/Caucasian | Lupus Class II | 109.43 | 0.06 |
| 33 | Female | White/Caucasian | Lupus Class III | 85.36 | 2.69 |
| 34 | Female | White/Caucasian | Lupus Class V | 109.48 | 2.19 |
| 35 | Female | #N/A | Lupus Class V | 111.95 | 1.56 |
| 36 | Female | Black/African American | Lupus Class V | 73.94 | 4.51 |
| 37 | Male | White/Caucasian | Diabetic nephropathy, hypertensive nephropathy | 66.24 | 1.49 |
| 38 | Male | | | | |
| 39 | Female | White/Caucasian | Diabetic nephropathy, hypertensive nephropathy, Tubulointerstitial disease | 19.52 | 9.11 |
| 40 | Female | White/Caucasian | Diabetic nephropathy | 15.75 | 1.39 |
| 41 | Male | White/Caucasian | Diabetic nephropathy | 56.05 | 0.91 |
| 42 | Female | White/Caucasian | Diabetic nephropathy | 44.68 | 6.74 |
| 43 | Female | Black/African American | Diabetic nephropathy | 64.23 | 1.01 |
| 44 | Male | White/Caucasian | Diabetic nephropathy | 18.29 | 6.49 |
| 45 | Female | White/Caucasian | Lupus Class V | 163.67 | 3.48 |
| 46 | Female | White/Caucasian | Lupus Class IV | 123.17 | 0.99 |
| 47 | Female | Asian/Asian American | Membranous Nephropathy | 180.4 | 3.19 |
| 48 | Male | White/Caucasian | IgA Nephropathy | 53.47 | 0.31 |
| 49 | Male | White/Caucasian | FSGS | 46.17 | 2.74 |
| 50 | Female | #N/A | IgA Nephropathy | 114.22 | 0.43 |
| 51 | Female | Asian/Asian American | Glomerular disease | 114 | 0.42 |
| 52 | Female | White/Caucasian | FSGS, IgA Nephropathy | 51.27 | 1.72 |
| 53 | Female | Black/African American | Membranous Nephropathy | 91.94 | 6.31 |
| 54 | Male | Black/African American | Diabetic nephropathy, Tubulointerstitial disease | 25.88 | 5.54 |
| 55 | Female | White/Caucasian | Diabetic nephropathy, Lupus Class II | 44.32 | 8.42 |
| 56 | Female | White/Caucasian | Diabetic nephropathy | 25.87 | 8.53 |
| 57 | Male | White/Caucasian | Diabetic nephropathy, FSGS | 48.72 | 0.53 |

**Table S2.** Confusion matrix and AUC for the eGFR at the biopsy for the top 7 features

| | | Prediction | |
|---|---|---|---|
| | | 0 | 1 |
| Actual | 0 | 34 | 2 |
| | 1 | 1 | 20 |
| **Accuracy** | | 0.95 | |
| **AUC** | | 0.96 | |
| **95% CI** | | 0.90 – 1.0 | |

**0:** eGFR ≥ 60, **1:** eGFR < 60, **CI**: confidence interval

**Table S3.** Confusion matrix and AUC for the eGFR in one year for the top 7 features

| | | Prediction | |
|---|---|---|---|
| | | 0 | 1 |
| Actual | 0 | 25 | 5 |
| | 1 | 4 | 23 |
| **Accuracy** | | 0.84 | |
| **AUC** | | 0.85 | |
| **95% CI** | | 0.76 – 0.95 | |

**0:** eGFR slope < 0, **1:** eGFR slope ≥ 0, **CI**: confidence interval

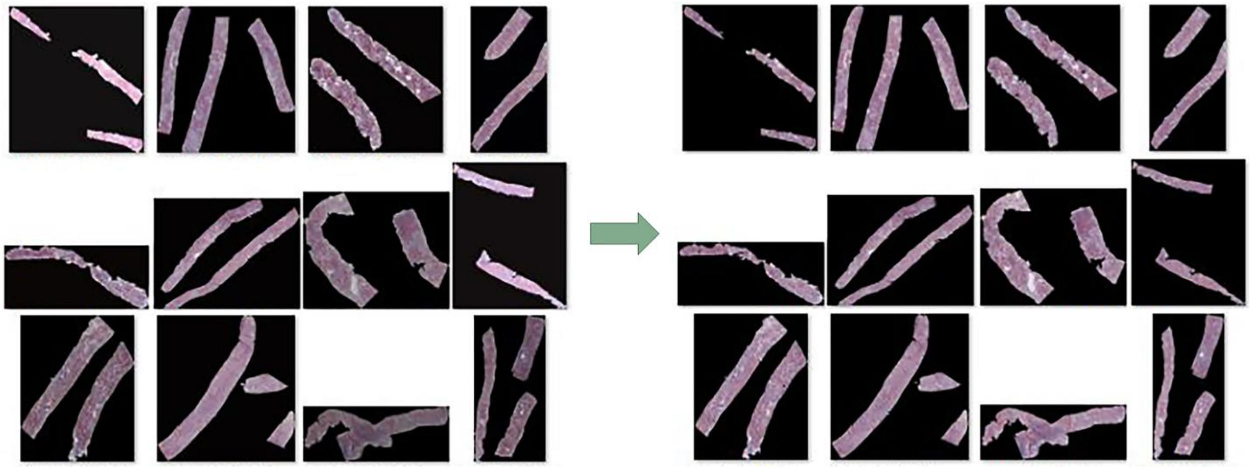

**Figure S1.** Stain color normalization. (A) Example images of biopsy samples. Multiple cortexes are combined in each case. To reduce the color and intensity variations present in the stained images, we computed the global mean and standard deviation of each channel in the Lab color space for the Reinhard color normalization for all data and used them as reference values to normalize our data. The figure shows Reinhard color normalization before (left) and after (right)[1].

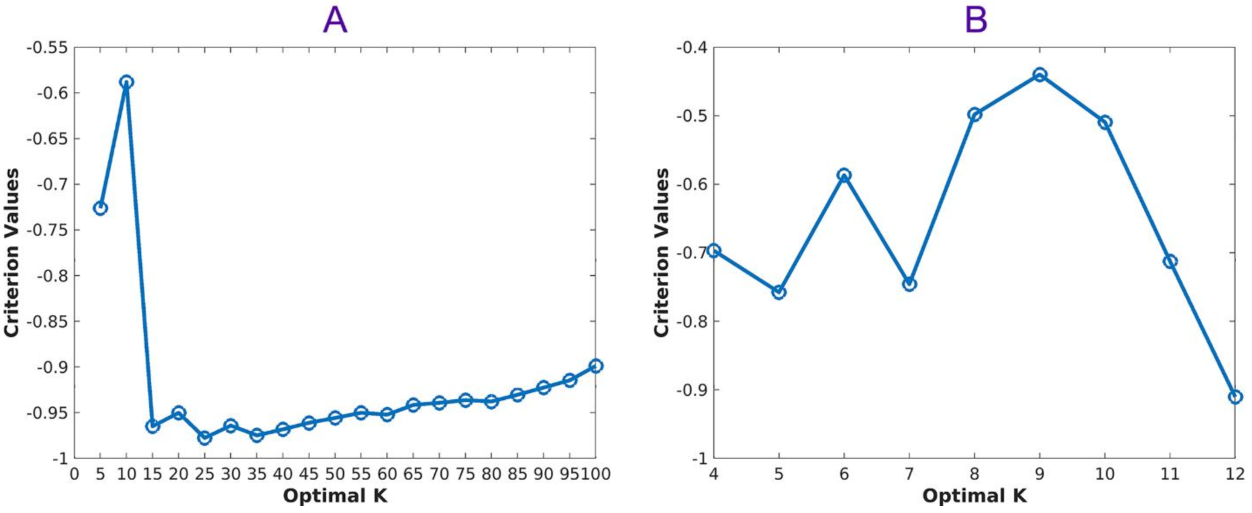


**Figure S2.** Determining an optimal number (K) of clusters using the Silhouette algorithm. (A) First, we run the algorithm every 5th point between 5 and 100 and then (B) run the algorithm between 4 and 12 to find the optimal K = 9 for the K means clustering[1].

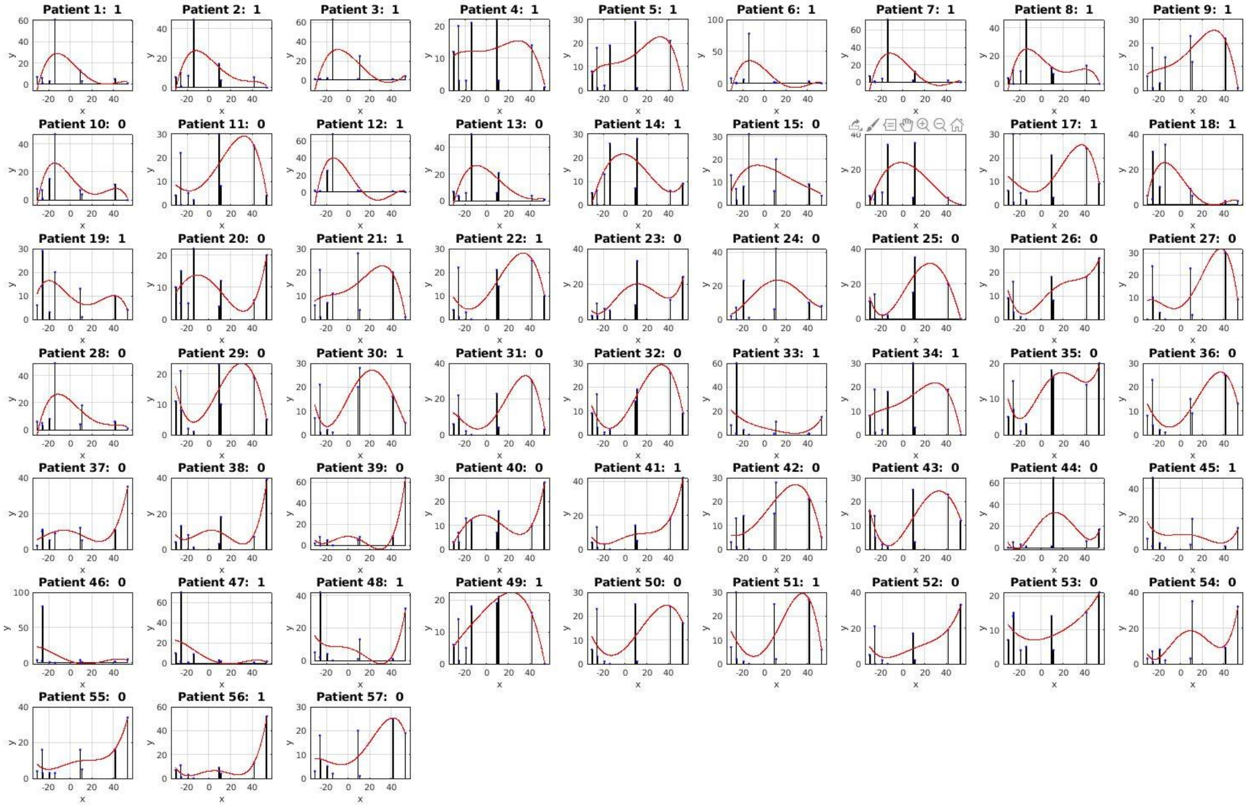


**Figure S3**. Frequency Histograms with the $4^{th}$ polynomial fittings for all 57 cases with labels (0 for eGFR ≥ 60 and 1 for eGFR < 60). The x-axis represents the distance between cluster groups obtained from MDS and y-axis represents the normalized frequencies of the clusters[1].

### References

1 Lee, J. *et al.* Unsupervised machine learning for identifying important visual features through bag-of-words using histopathology data from chronic kidney disease. *Sci Rep* **12**, 4832, doi:10.1038/s41598-022-08974-8 (2022).
